# Decreased GPR55 expression links B-cell activation and vascular remodelling in atherosclerosis in early rheumatoid arthritis patients

**DOI:** 10.1101/2025.03.06.25323490

**Authors:** Daniel Miranda-Prieto, Mercedes Alperi-López, Ángel I. Pérez-Álvarez, Sara Alonso-Castro, Ana Suárez, Javier Rodríguez-Carrio

## Abstract

**Objective:** inflammation and repair responses may be involved in atherosclerosis in rheumatoid arthritis (RA), although mechanisms are unknown. GPR55 is a cannabinoid receptor expressed in hematopoietic and stromal tissues, which has been implicated in atherosclerosis in mouse models, but evidence in humans is lacking. Our aim was to evaluate GPR55 expression in leukocyte populations in RA patients and their potential role in atherosclerosis.

**Methods:** GPR55 expression was quantified by flow cytometry in 63 untreated RA patients, 11 arthralgia individuals and 36 controls. Atherosclerosis was assessed by Doppler-ultrasound. Cytokines were measured by immunoassays, and serum proteomics were performed by a high-throughput targeted panel. In vitro cultures were performed with mononuclear cells from healthy donors.

**Results:** Decreased GPR55 expression in B-cells and monocytes was found in RA, whereas no differences were observed in arthralgia. Public datasets validated these findings. B-cell GPR55 expression was unrelated to clinical features, risk factors and atherosclerosis in RA, but exhibited divergent associations with leukocyte populations. GPR55 expression was associated with proinflammatory cytokines, and proteomic signatures related to vascular remodelling and B-cell responses in RA. These associations were dependent on the atherosclerosis status. LPS exposure in vitro decreased GPR55 expression in B-cells in a dose-dependent manner, which overlapped increasing CB86 expression.

**Conclusions:** Reduced GPR55 expression hallmarked B-cells and monocyte subsets in early RA. GPR55 expression was linked to B-cell activation-related pathways, presumably via T-cell independent mechanisms, and vascular remodelling. GPR55 may be a novel hub to understand the crosstalk between immune circuits and maladaptive responses in atherosclerosis.

## INTRODUCTION

Rheumatoid arthritis (RA) has been consistently linked to an increased risk of atherosclerosis occurrence and progression. This risk excess cannot be solely explained by traditional cardiovascular (CV) risk factors, and the role of inflammation has been proposed [1,2]. However, specific mechanisms and mediators are far from being clear. Atherosclerosis is a complex, chronic disease which involves different cell populations from different organs and tissues, and processes such as inflammation, remodelling and repair [3]. Thus, an intricate network of shared mechanisms and mediators ultimately orchestrate this multi-organ crosstalk.

The endocannabinoid system is an endogenous lipid-signalling system that regulates several pathways in nervous and peripheral tissues, ranging from pain, energy homeostasis, lipid metabolism to vascular tone and inflammation [4,5]. The endocannabinoid system relies on several endogenous lipid mediators, enzymes involved in endocannabinoid metabolism, and G protein-coupled receptors (GPCR) [6]. Classical cannabinoid (CB) receptors have been reported to trigger different effects on the immune response, with CB1 activation shown to be rather pro-inflammatory and CB2 promoting anti-inflammatory effects [7,8]. Recently, other GPCR sensitive to endocannabinoids have been described, including GPR55 [4]. GPR55 has been found to exhibit a low homology with classical CB receptors and lacks conventional cannabinoid binding pockets [9], hence suggesting different functions. In fact, GPR55 is thought to use different signalling pathways and be responsive to a range of ligands [10] when compared to classical CB receptors.

Novel evidence has demonstrated a pivotal role for GPR55 in atherosclerosis by shaping the B-cell compartment in knock-out animal models [11], hence underscoring the role of this receptor in the immune system. GPR55 may thus explain the anti-inflammatory effects of certain endocannabinoids [12] as well as account for the increased atherogenesis upon cannabinoid treatments [7]. However, data on the expression of GPR55 in human leukocytes is rather limited [13], and most evidence came from murine immune transcriptomic atlas and animal studies [14]. Thus, evidence on the role of GPR55 in human B-cells is lacking. Despite various GPCRs have been linked with immune responses, inflammation and clinical outcomes [15,16], whether GPR55 may be involved in human diseases remains to be explored. Interestingly, contemporary research has demonstrated that GPR55 modulation hold promise as a pharmacological target in metabolically active tissues to improve disease outcomes [17,18], and potentially atherosclerosis [19], in animal studies. Of note, the endocannabinoid system has been proven to participate in other pathological outcomes, including cardiometabolic risk [20]. Although the endocannabinoid system has been studied in RA by virtue of its implication in analgesia, inflammatory pain and neural-immune axis [21,22], its role in comorbidities, including CV disease and atherosclerosis, has been largely neglected.

Taken together, we hypothesized that GPR55 expression may be altered in RA patients, probably in association with atherosclerosis outcomes. Therefore, the main aims of the present study were (i) to evaluate the GPR55 expression in leukocytes from RA patients and healthy volunteers, (ii) to assess their associations with clinical and immunological features, and (iii) to characterize functional relevance and underlying causes of GPR55 expression in RA.

## MATERIAL AND METHODS

### Study participants

RA patients fulfilling the 2010 ACR/EULAR classification criteria [23]were recruited from the early arthritic clinic of the Department of Rheumatology at Hospital Universitario Central de Asturias (HUCA). RA patients were recruited at disease onset; therefore, they were not exposed to disease-modifying antirheumatic drugs (DMARDs) at the time of sampling. During the patients’ appointment, a complete medical examination was performed, including Disease Activity Score 28-joints (DAS28) and Health Assessment Questionnaire (HAQ) calculations. A group of individuals with clinically suspect arthralgia (CSA) [24] were also recruited from the same clinic. Additionally, healthy controls (HC) were recruited among unrelated, age- and sex-matched healthy volunteers from the same population.

A fasting blood sample was collected from all individuals by venepuncture in EDTA-containing tubes. Blood samples were immediately transferred to the laboratory and processed within less than 2 hours. Serum samples were stored at −80°C until experimental procedures. Conventional blood biochemical (including CRP and ESR measurements), lipid analyses, and complete blood counts were performed in all individuals. Traditional CV risk factors (dyslipidemia, hypertension, diabetes, smoking habits and obesity) in compliance with national guidelines were obtained from the medical records.

The study was approved by the local institutional review board (Comité de Ética de Investigación con Medicamentos del Principado de Asturias) in compliance with the Declaration of Helsinki (reference CEImPA 2021.126). All study subjects gave written informed consent.

### Vascular imaging

Subclinical atherosclerosis was assessed by Doppler ultrasound at the sonography laboratory (Department of Neurology, HUCA) by an experienced user blinded to the status of the study participants. All measures were performed online in B-mode using a Toshiba Aplio XG device (Toshiba American Medical Systems). The carotid intima-media thickness (cIMT) was measured at left and right common carotid arteries, as previously described [25], according to the “Mannheim Carotid Intima-Media Thickness Consensus (2004-2006)” [26]. Atherosclerotic plaque was established according to consensus definition [25]. Plaque risk was assessed by the ultrasound appearance of the plaques, and these were classified as “low” or “high” risk [27].

### Analysis of GPR55 expression

Peripheral Blood Mononuclear Cells (PBMCs) fractions were obtained by centrifugation (1800 rpm, 20 minutes) on density gradients (Lymphosep, Biowest, Germany). PBMCs (1·10^6^ cells/ml) were stained with anti-CD19 PerCP-Cy5,5 (Immunostep, Spain), anti-CD14 allophycocyanin (APC, Immunostep) and anti-GPR55 phycoerythrin (PE, BIOSS Antibodies, USA), or isotype controls (all from Immunostep) for 30 minutes at 4°C. Next, cells were washed twice with PBS and acquired in a BD FACS Canto II flow cytometer, using FACS Diva 2.6 for analysis. Lymphocytes and monocytes were gated according to their differential FSC/SSC features. CD19+ cells within the lymphocyte population were selected, and GPR55 expression was analyzed in a histogram for the PE channel to quantify GPR55+ B-cells. Similarly, monocyte subsets were assessed for GPR55-expressing CD14low and CD14high populations. Furthermore, GPR55 expression was also quantified by the mean fluorescence intensity (MFI) from gated populations (CD19+, CD14low, and CD14high), after subtracting MFI values from isotype controls.

### Assessment of cytokine levels

IL-6, TNF, IFNγ, IL-1b, IL-33, IL-23, IL-18, IL-17, IL-12, IL-10, and IL-8 serum levels were measured by a predefined multiplex assay (Human Inflammation Panel 1, LEGENDplex, BioLegend) in a BD FACS Canto II flow cytometer, according to the protocol provided by the manufacturer. The detection limits were 2.68 pg/ml, 2.90 pg/ml, 2.68 pg/ml, 3.17 pg/ml, 19.53 pg/ml, 4.39 pg/ml, 3.90 pg/ml, 0.73 pg/ml, 3.40 pg/ml, 3.66 pg/ml and 2.92 pg/ml, respectively. The serum levels of IFNα were quantified using a Cytometric Bead Array Flex Set (BD) in a BD FACS Canto II flow cytometer using FCAP Array v.1.0.1, following the manufacturer’s instructions. The detection limit was 1.25 pg/ml.

### Proteomic analyses

Serum proteomics were evaluated through a high-throughput analysis. A pre-defined panel of 92 protein hits related to CV (CV Panel II) was measured using the Proximity Extension Assay proprietary test from Olink (Olink Bioscience, Sweden) [28].

### In vitro cultures

PBMCs from healthy donors were isolated as previously described. PBMCs were cultured (37°C, 5% CO_2_) at a density of 2·10^6^ cells/ml in RPMI 1640 medium (Biowest) supplemented with 10% FCS (Biowest) and streptomycin/ampicillin (Sigma Merk, Belgium) (50 μg/ml and 250 μg/ml, respectively) under five LPS (Invitrogen, Germany) concentrations (0, 10, 100, 1000 and 5000 ng/ml) for 24, 48 or 72h.

After desired timepoints, cultured PBMCs from each condition were recovered from 48-well plates to analyze the GPR55 expression by flow cytometry. PBMCs were stained with anti-CD19 PerCP-Cy5,5 and anti-GPR55 PE antibodies as previously described. Moreover, 7-AAD PerCP-Cy-5,5 (Immunostep, Spain) was added to 100 µl PBMCs from each condition to monitor PBMCs viability.

### Statistical analyses

Variables were summarized as median (interquartile range) or n(%), depending on their distribution. Differences among groups were evaluated by Mann-Withney U or Kruskal-Wallis tests, as appropriate. Corrections for multiple comparison tests were performed by Dunn-Bonferroni. Correlations were assessed by Spearman ranks’ tests.

Gene expression datasets were downloaded from the publicly available NCBI Gene Expression Omnibus (GEO) repository [29]. GPR55 expression data was tested to be differentially regulated in the default patient groups using the GEO2R tool (GEOquery and limma R packages) and the corresponding FDR-adjusted p-value was calculated. Then, target data were downloaded and presented in graphs (analysis by conventional tests). Proteomic data were evaluated under STRING platform to evaluate protein-protein interactions as well as to identify pathway enrichment analyses using KEGG library [30].

A p-value <0.050 was considered as statistically significant. Statistical analyses were carried out under SPSS v. 27, R v.4.1.3 and GraphPad Prism 8.0.

## RESULTS

### GPR55 expression in B-cells and monocytes from RA patients

A total of 63 early RA patients, 11 CSA individuals and 36 sex- and age-matched HC were recruited for this study (Table 1). GPR55 expression was detected in the surface of B-cell and monocytes (both CD14high and CD14low subsets) from study participants by flow cytometry (Figure 1A). RA patients exhibited a lower GPR55 expression (measured as mean fluorescence intensity) in B-cells and monocyte subsets compared to HC (Figure 1B). Equivalent results were obtained when the frequency of GPR55+ cells was computed (CD19+: 6.81 (4.17) vs 12.62 (17.00)%, p=0.031; CD14high: 31.77 (17.25) vs 37.54 (18.04)%, p=0.076; and CD14low: 37.08 (15.77) vs 46.34 (23.28)%, p=0.002). No differences were observed between CSA individuals and HC in all subsets (all p>0.050).

**Figure 1:**
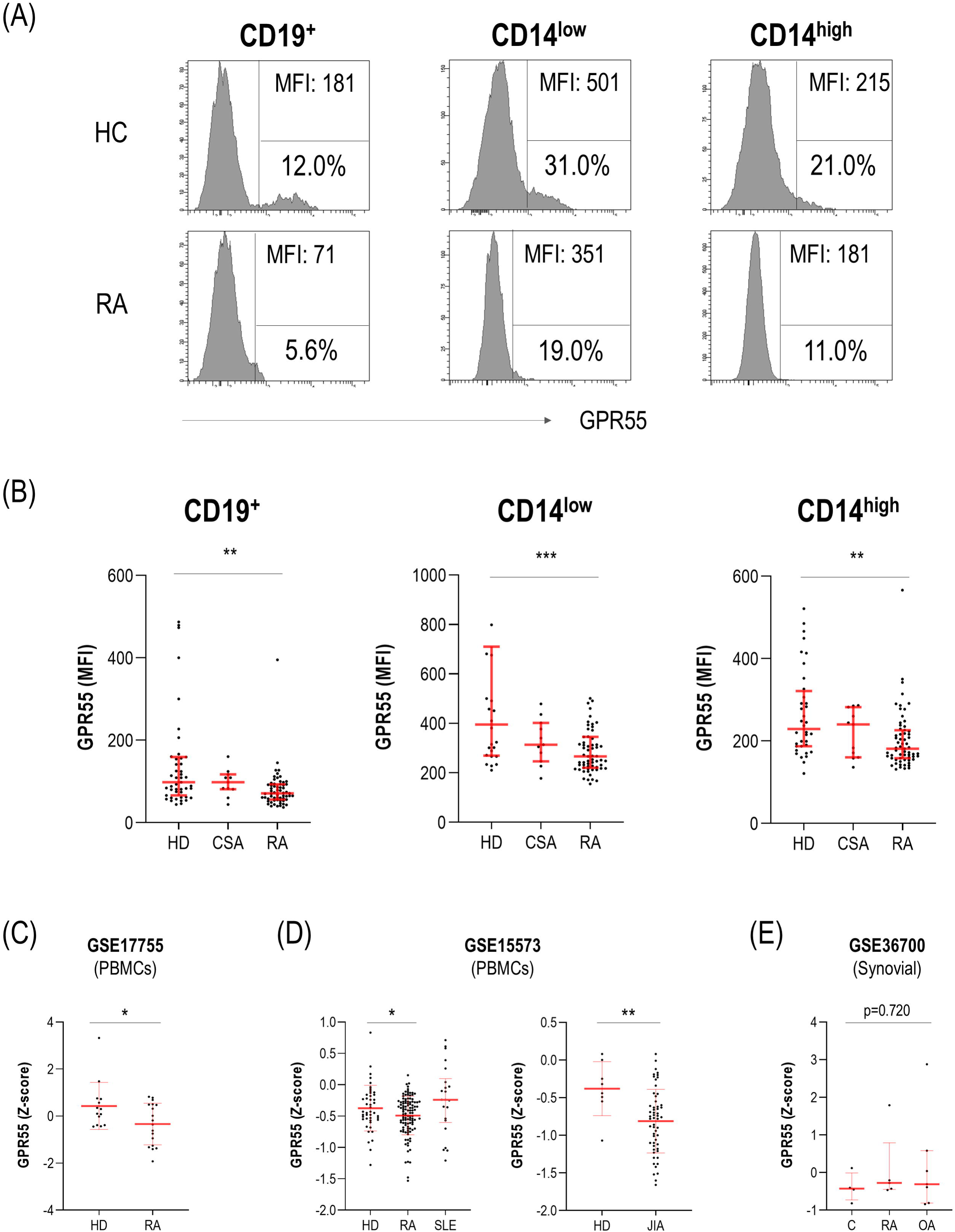
Expression of GPR55 in human leukocytes. (A) GPR55 expression was assessed by flow cytometry in CD19+, CD14low and CD14high cell subsets. Representative dot plots from a HC and a RA patient are shown. MFI (top) and frequency (5) of GPR55+ cells (bottom) within each subset is indicated. (B) GPR55 expression (MFI) was compared among HC, CSA and RA groups. GPR55 expression in gene expression datasets from PBMC (C-D) and synovial tissue (E) were analyzed to validate our findings. In scatter plots, each dot represents one individual. Bars represent 25th percentile, median and 75th percentile. Differences were assessed by Kruskal-Wallis or Mann-Withney with Dunn-Bonferroni tests for multiple comparisons. p-values correspond to those obtained in the multiple comparisons tests and are indicated as follows: * p<0.050, ** p<0.010 and *** p<0.001.

**Table 1:** Demographic and clinical features of study participants. Demographic and clinical features of study participants were summarized. Variables were expressed as median (interquartile range) or n(%), unless otherwise stated, according to the distribution of the variables.

|  | **HC**<br>**n=36** | **CSA**<br>**n=11** | **RA**<br>**n=63** |
| --- | --- | --- | --- |
| Age (years), mean (range) | 59.30<br>(38.30-81.00) | 49.77<br>(38.08-62.33) | 58.61<br>(30.33-83.67) |
| Gender (female/male) | 22/14 | 11/0 | 52/11 |
| ***Clinical features*** |  |  |  |
| Duration of symptoms (weeks) |  | 24.00 (40.00) | 18.00 (20.00) |
| Morning stiffness (minutes) |  | 30.00 (20.00) | 60.00 (105.00) |
| Tender joint count |  | 0.00 (0.00) | 6.00 (6.00) |
| Swollen joint count |  | 3.00 (3.00) | 9.00 (9.00) |
| ESR (mm/h) |  | 5.00 (8.00) | 24.00 (24.00) |
| CRP (mg/dl) |  | 0.20 (0.30) | 0.80 (2.10) |
| Patient global assessment (VAS<br>0-100) |  | 30.00 (40.00) | 70.00 (20.00) |
| DAS28 |  |  | 5.25 (1.68) |
| HAQ |  | 0.60 (0.60) | 1.12 (1.00) |
| Pain (VAS 0-10) |  | 5.00 (4.00) | 7.00 (2.00) |
| RF+, n(%) |  | 6 (54.5) | 44 (69.8) |
| ACPA+, n(%) |  | 5 (45.4) | 42 (66.6) |
| Shared epitope, n(%) |  | 3 (27.2) | 42 (66.6) |
| ***Traditional CV risk factors*** |  |  |  |
| Hypertension, n(%) |  | 1 (9.1) | 21 (33.3) |
| Diabetes, n(%) |  | 0 (0.0) | 5 (7.9) |
| Dyslipidemia, n(%) |  | 3 (27.3) | 19 (30.1) |
| Smoking, n(%) |  | 7 (63.6) | 26 (41.2) |
| Obesity (BMI>30 kg/m<sup>2</sup>), n(%) |  | 1 (9.1) | 22 (34.9) |
| Waist circumference (cm) |  | 90.00 (11.50) | 99.50 (20.25) |

Furthermore, data from GPR55 gene expression was extracted from publicly available microarray datasets from the GEO database in order to validate our findings. Three datasets containing relevant samples for our analysis were retrieved: 2 datasets from PBMCs studies in RA patients and 1 dataset including stromal (synovial) tissue. GSE17755 included gene expression data from 18 RA patients and 15 HC, thus revealing a GPR55 downregulation in the RA group (Figure 1C). GSE15573 included data from RA (n=112), SLE (n=22), juvenile idiopathic arthritis (JIA) (n=57), and healthy donors (n=53). GPR55 was found to be differentially expressed by GEO2R (adjusted p=2.05·10^-10^ among groups). Subgroup analysis revealed diminished GPR55 expression in RA and JIA groups compared to control groups, whereas no differences were noted in SLE patients (Figure 1D). Finally, results from GSE36700 containing synovial tissue samples from 7 RA patients, 5 osteoarthritis (OA) patients and 5 patients with microcrystalline arthritis revealed no differences in GPR55 expression at the local tissue level (Figure 1E).

Taken together, these results confirm that GPR55 is expressed in leukocyte subsets from human subjects, namely B-cells and monocytes. RA patients were hallmarked by a decreased GPR55 expression, both at protein and gene expression levels.

### GPR55 expression in B-cells is unrelated to clinical features and atherosclerosis status in RA patients

Next, associations between GPR55 expression and clinical features, traditional CV risk factors and subclinical atherosclerosis were evaluated.

GPR55 expression was unrelated to clinical features, including disease activity, joint counts or symptom duration in RA patients, regardless of the cell subset analyzed (Supplementary Table 1). Equivalent results were obtained for CSA individuals. Similarly, GPR55 expression was not associated with traditional CV risk factor overall. However, smoking was associated with a lower GPR55 expression in B-cells (p=0.047), whereas no effect was observed in monocytes (CD14high: p=0.175, and CD14low: p=0.234). Interestingly, this effect was dose-dependent, since a correlation with the number of cigarettes/day was retrieved (r=-0.249, p=0.039), and restricted to patients carrying the shared epitope (p<0.050, and r=-0.375, p=0.027; respectively), while being absent in those without these alleles (p=0.592; and r=-0.165, p=0.431; respectively).

GPR55 expression in B-cells showed opposed associations with the frequency of leukocyte subsets (neutrophils: r=0.383, p=0.002; and lymphocytes: r=-0.387, p=0.003) in RA patients. Furthermore, GPR55 expression in B-cells negatively paralleled the size of the CD19+ population (% CD19+: r=-0.568, p<0.001; absolute counts: r=-0.646, p<0.001), and may be attributed to the frequency of GPR55-B-cells (r=-0.756, p<0.001), which remained associated with GPR55 expression even after correcting for total B-cell frequency (r=-0.331, p<0.001). Of note, no association was found with CD3+ frequency (r=-0.179, p=0.164). Equivalent results were also observed in CSA individuals (CD19+: r=-0.630, p=0.038; and CD3+: r=0.110, p=0.748). No associations were found with GPR55 expression on monocytes.

Finally, neither atherosclerosis occurrence nor cIMT (Supplementary Table 2) were related to GPR55 expression in B-cells in RA patients (p=0.146, and r=0.027, p=0.841; respectively). Plaque burden or presence of high risk plaques were unrelated to GPR55 expression (r=-0.039, p=0.750; and p=0.205; respectively). Similar findings were observed for that of monocyte expression (all p>0.050).

All these results suggest that reduced GPR55 expression in RA is unrelated to clinical features, traditional CV risk factors or atherosclerosis burden. However, divergent associations with leukocyte populations were noted, pointing to an effect within the B-cell compartment.

### GPR55 expression in B-cells was associated with vascular remodelling and B-cell proteomic signatures in RA patients

The analysis of serum cytokines revealed different trends in their association with GPR55 expression depending on the cellular subsets (Table 2). Interestingly, GPR55 expression was correlated with several proinflammatory cytokines (IL-6, IL-8, and IL-18) in RA patients, while a different picture was retrieved for monocyte subsets. No associations were observed in CSA or HC groups (all p>0.050).

**Table 2:** GPR55 expression and proinflammatory cytokines. The associations between GPR55 expression in different cellular subsets and serum levels of proinflammatory cytokines were analysed by Spearman’s rank tests in RA patients. Correlation coefficients (r) and p-values are shown. Those reaching statistical significance are highlighted in bold.

|  | GPR55 expression |  |  |
| --- | --- | --- | --- |
|  | CD19+ | CD14low | CD14high |
| IL-6 | **r=0.236, p=0.062** | r=0.162, p=0.205 | r=0.090, p=0.483 |
| TNF | r=0.110, p=0.390 | r=0.192, p=0.131 | r=0.106, p=0.409 |
| IFNγ | r=0.165, p=0.196 | r=0.135, p=0.291 | r=0.039, p=0.762 |
| IFNα | r=0.092, p=0.473 | r=0.082, p=0.523 | r=-0.058, p=0.651 |
| IL-1β | r=0.097, p=0.451 | r=0.101, p=0.429 | r=-0.052, p=0.684 |
| IL-33 | r=0.067, p=0.603 | r=0.024, p=0.849 | r=-0.096, p=0.456 |
| IL-23 | r=0.162, p=0.206 | r=0.090, p=0.485 | r=0.076, p=0.552 |
| IL-18 | **r=0.410, p<0.001** | **r=0.318, p=0.011** | r=0.164, p=0.198 |
| IL-17 | r=0.218, p=0.086 | r=0.042, p=0.744 | r=0.036, p=0.778 |
| IL-12 | r=0.086, p=0.505 | r=0.010, p=0.937 | r=-0.089, p=0.488 |
| IL-10 | r=0.178, p=0.163 | r=0.057, p=0.659 | r=-0.013, p=0.922 |
| IL-8 | **r=0.317, p=0.011** | r=0.226, p=0.074 | **r=0.266, p=0.035** |

The assessment of serum proteomics provided similar results. Correlation analyses revealed that GPR55 expression in B-cells showed associations with a total of 15 protein hits, whereas 4 did with that of CD14high monocytes, and no associations were observed for the CD14low subset. Equivalent results were obtained when GPR55+ frequency was used. Overall, proteins correlated with GPR55 expression in B-cells were related to matrix turnover and remodelling (PGF, CXCL1, IL-18, SERPINA12, MMP7, MMP12, ADAMTS13, and THBS2) and B-cell activation (SLAMF7, IL-6, VSIG2, and TNFRSF13B). Further analyses revealed a significant protein-protein interaction enrichment (p=5.99·10^-11^) using the STRING platform (Figure 2A). Pathway annotation using KEGG mapper identified pathways related to humoral/type 2 adaptive responses, cytokine and chemokine production, neuroinflammation, and foam cell differentiation (Figure 2A). Of note, these correlations were dependent on the atherosclerosis status, being present only in patients with atherosclerosis and absent in the atherosclerosis-free group (data not shown). In fact, subgroup analysis of patients with atherosclerosis mostly recapitulates the protein hits (n=11, protein-protein interaction enrichment: p=8.96·10^-7^) and signatures observed at the whole group level (Figure 2B), hence suggesting that correlations at group level were mostly driven by the subgroup of patients with atherosclerosis. Interestingly, cytokine production and cellular response to LPS pathways predominated in this analysis.

**Figure 2:**
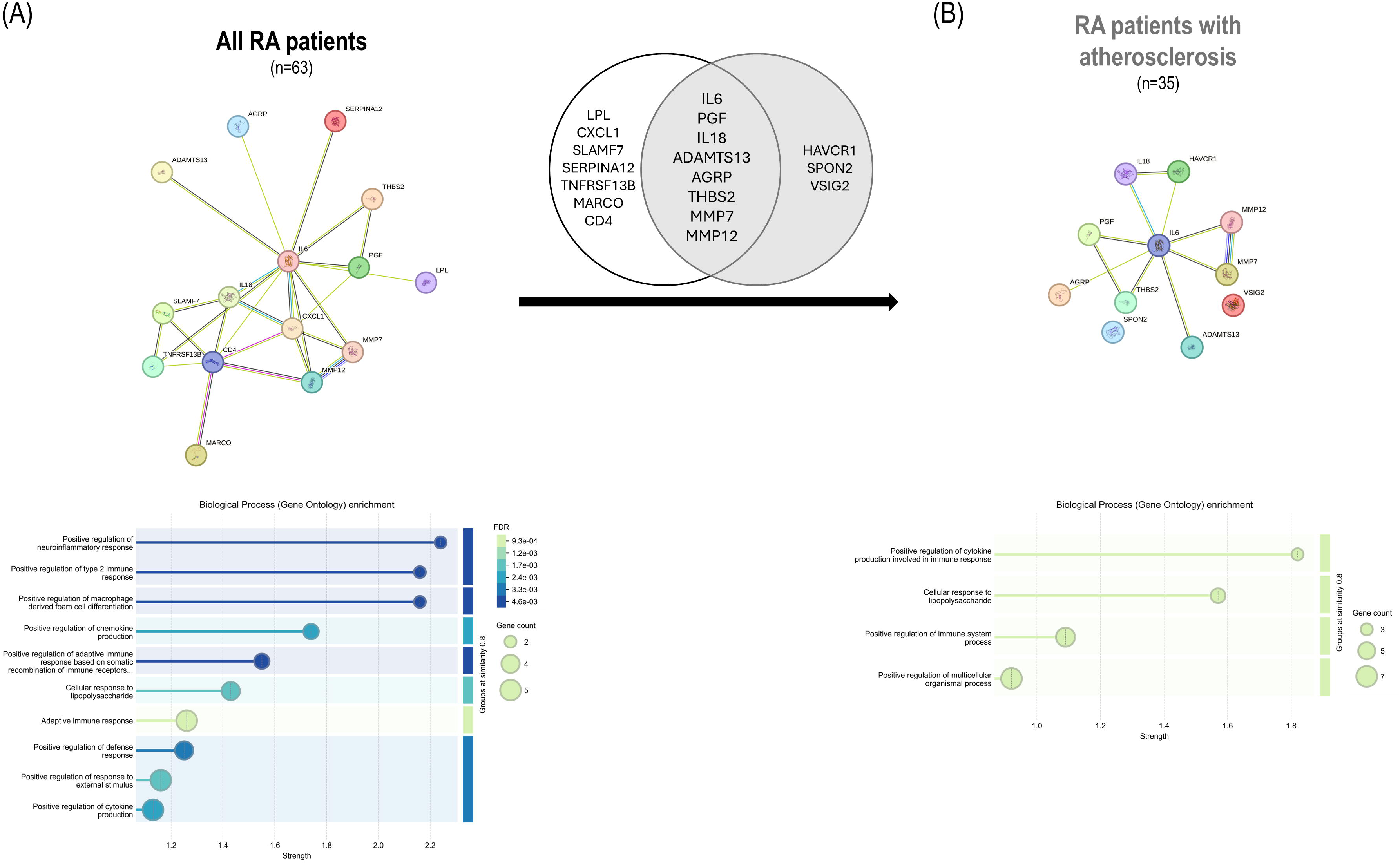
GPR55 and serum proteomics. (A) GPR55 expression in B-cells was associated with a number of protein hits in RA patients. These proteins exhibited a high degree of interaction (network graph, top) and informed functional pathways (biological process enrichment graph, bottom). Strength, FDR and gene/protein counts are indicated for each pathway. These associations in the whole RA group showed a high degree of overlap with those retrieved in RA patients with atherosclerosis (B).

All these results highlight distinct associations of GPR55 expression depending on cellular subsets. GPR55 expression in B-cells was related to proinflammatory and B-cell responses as well as with vascular remodelling pathways in an atherosclerosis-restricted manner.

### LPS induced a dose-dependent GPR55 downregulation in human B-cells

Next, in order to gain insight into the GPR55 downregulation in B-cells, in vitro experiments were carried out with PBMCs from healthy volunteers exposed to growing concentrations of LPS along different timepoints.

In vitro assays demonstrated that LPS induced a GPR55 downregulation in B-cells (Figure 3A). Although this effect was more evident at higher concentrations (1000-5000 ng/ml), certain effect was also observed at lower levels (10-100 ng/ml) (Figure 3A), and statistical analyses revealed a dose-dependent effect along the whole range (r=-0.832, p for trend < 0.001). LPS-mediated GPR55 reduction overlapped with increasing CD86 expression, hence linking GPR55 downregulation with B-cell activation, also arising at lower LPS concentrations (Figure 3B). Lowered GPR55 expression could not be attributed to a decrease of the total B-cell population, as it was unaffected by LPS exposure (Figure 3C). Similarly, a potential effect on viability can be excluded, and no toxicity was registered even at higher concentrations (frequency of 7-AAD+ cells within the live gate ranged between 2-4%), thus ruling out a potential confounding effect. Finally, these effects occurred early (24h), but they were stable along longer timepoints (48-72h) (Supplementary Figure 1).

**Figure 3:**
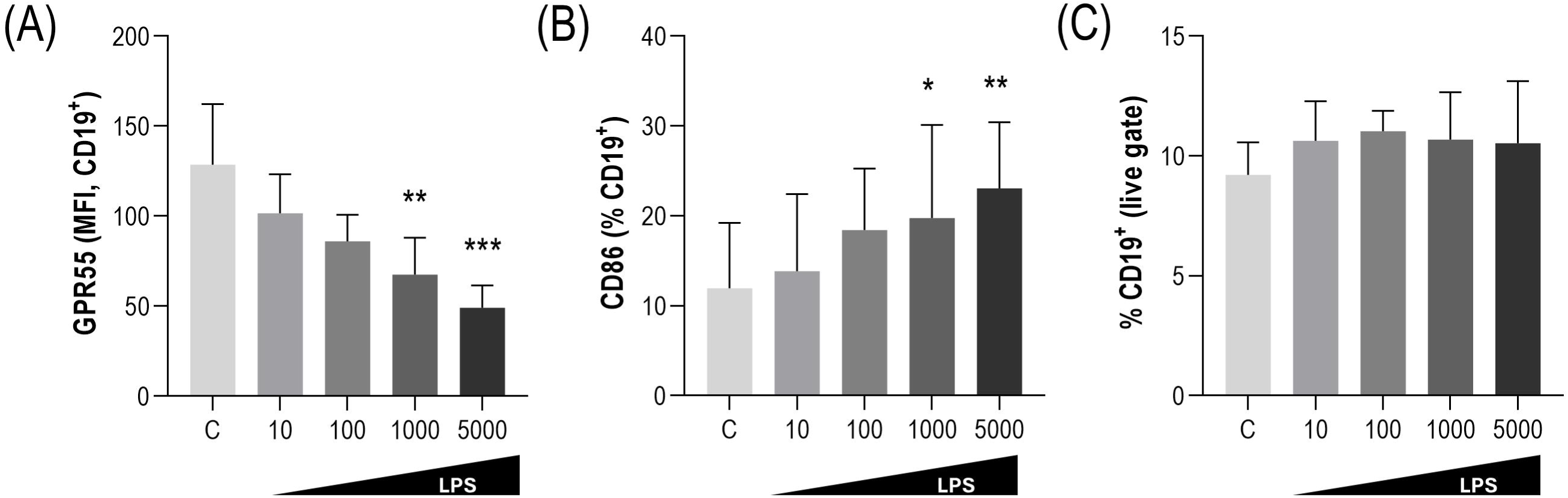
Effect of LPS on GPR55 expression. (A) The effect of LPS on GPR55 expression in B-cells at 24h was evaluated in vitro (n=6 for each condition). The effect on CD86 expression (B) and CD19+ total population (C) was also analyzed by flow cytometry. Bars represent median values and interquartile range. Differences were assessed by Kruskal-Wallis with Dunn-Bonferroni tests for multiple comparisons. p-values correspond to those obtained in the multiple comparisons tests and are indicated as follows: * p<0.050, ** p<0.010 and *** p<0.001.

Taken together, these results confirm that GPR55 expression in B-cells can be decreased in response to LPS exposure in a dose-dependent and sustained manner, thus suggesting an overlap between GPR55 downregulation and B-cell activation, presumably via T-cell independent mechanisms. Evidence from proteomic analyses support these findings.

## DISCUSSION

The involvement of B-cells in atherosclerosis has gained attention in recent years, although mechanistic underpinnings are yet to be clarified. The results herein reported demonstrate an altered expression of GPR55 in the early phase of RA, also in relation with inflammatory circuits, B-cell activation and remodelling responses. These results were independently validated in transcriptomic datasets and in vitro assays. To the best of our knowledge, this is the first description of GPR55 expression in patients with rheumatic and musculoskeletal diseases (RMDs).

Our findings demonstrate not only that GPR55 is expressed in leukocyte subsets from human populations, but also that altered GPR55 expression could be linked to human diseases, namely RA. These results were validated at the gene expression levels using publicly available datasets, also highlighting differences between cellular subsets (stromal vs hematopoietic). GPR55 expression in RA patients was unrelated to clinical features, including patient reported outcomes related to nociception, such as HAQ or pain scales. This finding emphasized the differences observed between GPR55 and other cannabinoid receptors [9]. Interestingly, evidence from cytokine analyses, proteomic signatures and in vitro experiments revealed associations between GPR55 expression, especially in B-cells, and inflammatory circuits. First, correlation analyses confirmed that GPR55 expression was associated with a number of pro-inflammatory mediators. Moreover, positive regulation of cytokine production was also retrieved after pathway annotation. Interestingly, distinct correlation profiles were noted between monocyte and B-cell subsets, thus pointing to cell context-specific roles or regulation. In fact, divergent patterns were also observed with lymphocytes and neutrophils counts. Furthermore, LPS exposure in vitro led to a dose-dependent GPR55 downregulation in B-cells. Equivalent results were obtained in neurons in an animal model, where the LPS challenge was also associated with a proinflammatory response [31]. LPS is a well-known stimulant of B-cells, hence prompting activation, proliferation and differentiation [32]. This may suggest a link between GPR55 expression and B-cell activation. Our findings support this idea, as LPS-induced GPR55 downregulation overlapped with increasing B-cell activation in vitro. Furthermore, the analysis of functional proteomic pathways also reinforced a potential connection with B-cell activation programmes. Recent evidence has reported that GPR55 deficiency may be connected with an altered IL-4 signalling [33], also related to B-cell differentiation. Transcriptomic and mitochondrial content analyses reinforced the involvement of GPR55 signalling in regulating B-cell activation and differentiation through a pleiotropic effect [11,34]. On the other hand, GPR55 signalling has been also reported to inhibit NFkB (reviewed in [35]), a master regulator of B-cell activation. Thus, it is tempting to speculate that reduced GPR55 signalling due to diminished GPR55 expression could in turn lead to an enhanced B-cell activation, thus creating a positive feedback loop. This may be of particular relevance under chronic stimulation, such as in autoimmunity. The negative association between GPR55 expression in B-cells and B-cell frequency in RA patients support this notion. Moreover, the positive association between GPR55 expression and neutrophil-to-lymphocyte ratio also aligns with a context of chronic stimulation. In fact, prolonged GPR55 stimulation has been reported to downregulated expression [36]. Collectively, our findings seem to suggest that GPR55 on monocytes and B-cells may not be related to pain, but rather to inflammation and B-cell activation. Decreased GPR55 may thus account for the B-cell overactivation observed in RA, hence pointing to a potential role as an actionable target or biomarker to guide therapeutic interventions.

In addition to immune circuits, GPR55 expression was also associated with several matrix metalloproteinases and other mediators such as IL-18, CXCL1 or THBS2, which are involved in vascular and extracellular matrix turnover in response to different stimuli. Pathway analysis from our proteomic platform validated this finding. These results are in line with the existing literature, as altered or blocked GPR55 signalling have been related to impaired remodelling upon experimental myocardial infarction [37], ventricular dysfunction [33] or cardiomyocyte hypertrophy [38]. Moreover, blocked GPR55 signalling has been linked to increased MMP-9 expression under inflammation [39,40]. Furthermore, GPR55 knock-down blunted angiogenesis and endothelial functions, including growth factors production, in vitro [41]. The findings herein presented are relevant in a twofold manner. First, we set the role of B-cells in the centre of this altered remodelling response. Whereas most of the existing evidence came from broad GPR55 deletion in animal models, our results are suggestive of a specific effect of the B-cell subset. This may explain why mice deficient in GPR55 in the haematopoietic system exhibited an aberrant remodelling response upon myocardial infarction, to some extent recapitulating that of cardiac-specific knock-out mice [37], despite stromal compartment was unaffected in the latter. Second, our findings expand previous evidence on the role of B-cell specific GPR55 deficient mice in atherosclerosis development. Lack of GPR55 had been reported to trigger B-cell hyperactivation [11], which is also in line with our findings. However, effects beyond B-cell compartment to account for vascular outcomes were largely unclear. Our study points to the involvement of remodelling and reparative processes in this scenario. GPR55 deficiency has been observed to modify the activity of a number of kinases, which relate to several cellular processes such as CREBP and mTORC2 signalling, response to oxidative stress, or even lipid metabolism [33]. Whether these kinases could also directly activate vascular remodelling, or those events may indirectly trigger this phenomenon is conceivable. Furthermore, altered tissue repair and matrix remodelling may be also related to limited GPR55 signalling, in a PPARg-mediated effect (reviewed in [35]). Functional studies are needed to elucidate the mechanisms underlying this crosstalk, also in order to identify therapeutic targets.

Experimental evidence has highlighted an association between GPR55 and vascular outcomes in animal models, including atherosclerosis. The association between GPR55 downregulation and B-cell activation may be in line with this protective effect on atherosclerosis. Surprisingly, our study failed to show a direct association between diminished GPR55 expression and atherosclerosis burden in RA patients. A number of remarks should be considered in this regard. Current evidence came from pre-clinical research using GPR55 knock-out models or strong pharmacological inhibition assays. Nevertheless, our study assessed the GPR55 expression at specific cellular subset level in real-world patient populations. Third, the nature of the association with vascular outcomes in the literature is intriguing. GPR55 deficiency has been linked to plaque size and phenotype, but not to atherosclerosis occurrence itself [11]. Similarly, infarct occurrence and size were unaffected by GPR55 status [37], although it led to an altered subsequent remodelling response. Equivalent results were observed for cardiomyocytes or left ventricular hypertrophy [33,38]. Furthermore, GPR55 antagonization did not affect plaque size or composition under different atherogenic conditions, but augmented plaque stability and infiltration in a CXCL1-dependent manner [39]. Taken together, these studies seem to rule out a direct, causative effect of decreased GPR55 on the occurrence of vascular outcomes. Alternatively, this evidence seems to suggest that diminished GPR55 may create a microenvironment more prone to vascular outcomes via altered remodelling, rather than a vascular insult itself. Experimental evidence also demonstrated that GPR55 led to decreased insulin resistance [42] or physical activity [43], which are also known to enhance, but not necessarily trigger, vascular outcomes. Additional hits may explain the occurrence of vascular events in certain individuals but not in others (Figure 4). This conceptual model aligns with the associations with reparative processes and/or maladaptive responses elsewhere described [33,37], also intertwining inflammation. It also supports the hypothesis that GPR55 signalling is required for the maintenance of physiological immune homeostasis under stress conditions [37]. Furthermore, this model also reconciles our findings with the existing literature, since the associations between GPR55 and remodelling, B-cell signatures, and inflammatory milieu were only observed in patients with atherosclerosis while being absent in their atherosclerosis-free counterparts. In fact, response to LPS proteomic signature predominated in patients with atherosclerosis, but not in those without atherosclerosis. This could be in line with the association between systemic LPS exposure and atherosclerosis occurrence in humans [44,45]. Moreover, GPCRs have been linked to intestinal barrier permeability [46], and GPR55 has been linked to intestinal inflammation [47], both being major sources of systemic LPS. However, the involvement of the B-cell subset in this regard remains unexplored. Taken together, all these lines of evidence help to understand how the atheroprotective role of B-cells depends on GPR55.

**Figure 4:**
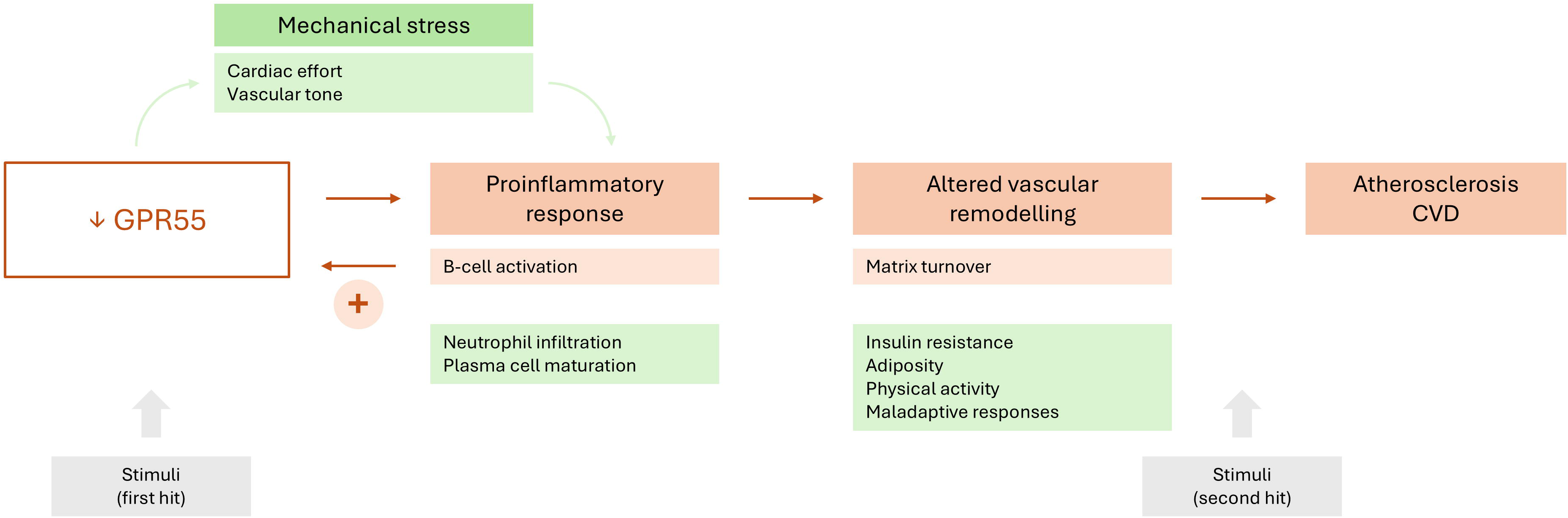
Association between GPR55 expression and atherosclerosis. Proposed model to explain the connection between GPR55 expression and atherosclerosis development. Certain hits, such as LPS exposure in this study, can lead to decreased GPR55 expression. This feature may directly cause a proinflammatory response affecting a number of immune cell subsets, including B-cells. Decreased GPR55 expression can also lead to proinflammatory response via mechanical stress, as observed in the heart. This proinflammatory response can in turn cause a lower GPR55 expression itself. Inflammation can also cause altered vascular remodelling, including matrix turnover and maladaptive responses, which enhance CV risk. Certain stimuli may act as ‘second hits’, hence triggering atherosclerosis or CVD occurrence. Orange squares represent evidence obtained in the present study, whereas evidence from the literature is summarized in green squares.

In conclusion, reduced GPR55 expression hallmarked monocytes and B-cell subsets in early arthritis, whereas no differences were found in the preclinical stage. GPR55 expression was linked to B-cell activation-related pathways, especially in patients with atherosclerosis, and presumably via T-cell-independent mechanisms. These findings open new avenues for treatment, aimed at reverting diminished GPR55 signalling, and may also help to understand the therapeutic outcomes upon certain drugs, as decreased GPR55 expression in certain conditions may limit beneficial effects. This may be of particular relevance as deficient GPR55 expression has been related to poor or delayed reparative responses [37], and increased adipogenic differentiation [48]. Moreover, reduced GPR55 may account for the enhanced neutrophil infiltration and activation in atherogenesis [39]. Of note, targeted GPR55 stimulation has been proposed to beneficially impact clinical outcomes upon infarction occurrence [37]. Furthermore, our findings strengthen the crosstalk between neuroendocrine, namely the endocannabinoid system, and immune circuits. Although targeting endocannabinoid system holds promise to block pain, inflammation, and joint destruction in RMDs, data is scarce and mostly limited to conventional CB receptors [49,50]. Thus, this study gained understanding towards novel mechanisms and mediators underlying this crosstalk. Emerging data from animal models also suggests a positive effect on atherosclerosis prevention [51], which may also support this notion. Moreover, our findings provide novel understanding towards the role of B-cells in atherosclerosis, especially in autoimmunity. Finally, the results herein presented expand the knowledge of non-conventional lipids in RA and atherosclerosis, as GPR55 signalling mostly rely on complex, arachidonic-derived lipid species such as lysophosphatidylinositol, palmitoylethanolamide or anandamide. Although lipoproteins have been largely studied, especially in the context of cardiovascular risk excess, other lipid compartments have been neglected. However, emerging evidence strengthen their role in recent years [52–54]. Taken together, GPR55 may be a novel hub to understand the crosstalk between immune responses and maladaptive remodelling in atherosclerosis.

## Author contributions

All authors were involved in drafting the manuscript or revising it critically for important intellectual content and all the authors gave their approval of the final version of the manuscript to be published.

Study conception and design: JRC, AS

Acquisition of data: DM-P, MAL, AIPA, SAC, AS, JRC

Analysis and interpretation of data: DM-P, JRC, MAL, AS

## Data availability statement

All data produced in the present work are contained in the manuscript.

## ACKNOWLEDGEMENTS

The authors would like to thank Ms. Marta García Boto (Department of Rheumatology, HUCA) for her assistance in the collection of blood samples, and the “Liga Reumatológica Asturiana” for their support.

