## Supplementary figures and images for "Decreased GPR55 expression links B-cell activation and vascular remodelling in atherosclerosis in early rheumatoid arthritis patients"

### Supplementary Figure 1

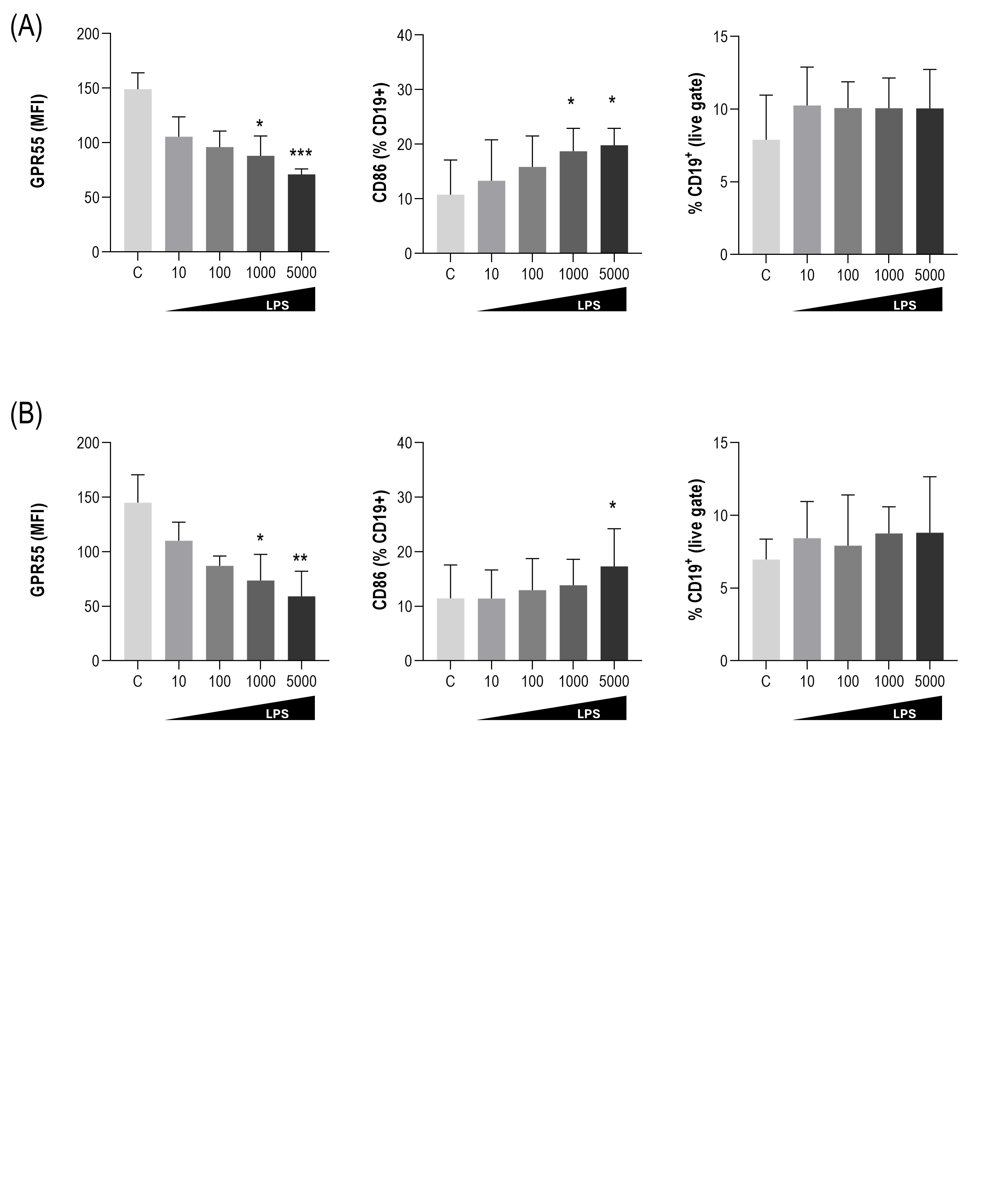
