## Supplementary Material for "Decreased GPR55 expression links B-cell activation and vascular remodelling in atherosclerosis in early rheumatoid arthritis patients"

**Supplementary Table 1**

**Supplementary Table 2**

**Supplementary Figure 1**

**Supplementary Table 1: GPR55 expression and clinical features.** The associations between GPR55 expression in different cellular subsets and serum levels of proinflammatory cytokines were analysed by Spearman’s rank tests in RA patients. Correlation coefficients (r) and p-values are shown. Those reaching statistical significance are highlighted in bold.

|  | **RA** | | |  | **CSA** |  |
| --- | --- | --- | --- | --- | --- | --- |
|  | **CD19+** | **CD14low** | **CD14high** | **CD19+** | **CD14low** | **CD14high** |
| ***Clinical features*** |  |  |  |  |  |  |
| Duration of symptoms | r=-0.028, p=0,836 | r=0.013, p=0.924 | r=0.079, p=0.558 | r=-0.144, p=0.674 | r=0.060, p=0.861 | r=-0.263, p=0.435 |
| Morning stiffness | r=0.154, p=0.229 | r=0.152, p=0.233 | r=0.085, p=0.508 | r=0.463, p=0.152 | r=0.379, p=0.251 | r=0.439, p=0.176 |
| Tender joint count | r=0.001, p=0.996 | r=0.087, p=0.495 | r=0.110, p=0.392 | r=-0.217, p=0.522 | r=-0.013, p=0.969 | r=0.067, p=0.844 |
| Swollen joint count | r=-0.027, p=0.832 | r=0.159, p=0.214 | r=0.138, p=0.282 | r=0.260, p=0.440 | r=0.277, p=0.409 | r=0.434, p=0.182 |
| ESR | r=0.089, p=0.488 | r=0.114, p=0.372 | r=0.184, p=0.149 | r=-0.139, p=0.685 | r=-0.179, p=0.598 | r=0.069, p=0.840 |
| CRP | r=0.131, p=0.306 | r=0.117, p=0.362 | r=0.141, p=0.272 | r=0.208, p=0.538 | r=0.354, p=0.285 | r=0.298, p=0.373 |
| DAS28 | r=0.029, p=0.821 | r=0.165, p=0.200 | r=0.177, p=0.168 | r=-0.068, p=0.841 | r=-0.009, p=0.979 | r=0.245, p=0.467 |
| HAQ | r=-0.113, p=0.384 | r=0.082, p=0.527 | r=-0.014, p=0.915 | r=-0.287, p=0.392 | r=-0.092, p=0.788 | r=-0.065, p=0.851 |
| Pain | r=-0.077, p=0.552 | r=-0.064, p=0.623 | r=0.031, p=0.811 | r=0.387, p=0.240 | **r=0.664, p=0.026** | r=0.538, p=0.088 |
| RF | p=0.134 | p=0.271 | p=0.496 | p=0.429 | p=0.247 | p=0.792 |
| ACPA | p=0.265 | p=0.614 | p=0.898 | p=0.792 | p=0.082 | p=0.429 |
| ***Traditional CV risk factors*** |  |  |  |  |  |  |
| Hypertension | p=0.498 | p=0.804 | p=0.965 | p=0.182 | p=0.727 | p=0.364 |
| Diabetes | p=0.612 | p=0.892 | p=0.278 |  |  |  |
| Dyslipidemia | p=0.225 | p=0.397 | p=0.222 | p=0.133 | p=0.921 | p=0.497 |
| Smoking | **p=0.047** | p=0.234 | p=0.175 | p=0.412 | p=0.412 | p=0.315 |
| Obesity | p=0.950 | p=0.438 | p=0.456 | p=0.400 | p=0.800 | p=0.800 |
| Waist circumference | r=0.100, p=0.948 | r=-0.021, p=0.884 | r=-0.050, p=0.732 | r=0.800, p=0.200 | r=0.200, p=0.800 | r=0.200, p=0.800 |

**Supplementary Table 2: Atherosclerosis features in RA and CSA groups.** Atherosclerosis features were summarized as indicated. The frequency of high risk plaques was calculated within the population of patients with plaque.

|  | **CSA** | **RA** |
| --- | --- | --- |
| ***Atherosclerosis*** |  |  |
| Presence of atherosclerosis, n(%) | 3 (27.2) | 35 (55.5) |
| Number of plaques, mean±SD | 0.50±0.97 | 0.98±1.07 |
| Plaque high risk, n(%) | 0 (0.0) | 13 (20.6) |
| cIMT, mean±SD | 0.57±0.09 | 0.67±0.16 |

**Supplementary Figure 1: Effect of LPS on GPR55 expression.** Effect of LPS on GPR55 and CD86 expression and CD19+ total population in vitro (n=6 for each condition) at 48h (A) and 72h (B). Bars represent median values and interquartile range. Differences were assessed by Kruskal-Wallis with Dunn-Bonferroni tests for multiple comparisons. p-values correspond to those obtained in the multiple comparisons tests and are indicated as follows: * p<0.050, ** p<0.010 and *** p<0.001.
